# Colonoscopy Complications in Persons with Spinal Cord Injury - worth the risk?

**DOI:** 10.64898/2025.12.19.25342693

**Authors:** Michelle Trbovich, Abigail McLaughlin, Christina Anthony, Wouker Koek, Alfredo Camero, Lilian Gowen, Keith Burau

## Abstract

**Objective:** Prevalence of anemia in persons with spinal cord injury is higher than their able-bodied counterparts at 50-80%. The etiology is multifactorial but likely related to chronic whole-body inflammation. Chronic anemia is often an indication of gastrointestinal blood loss leading to a diagnostic colonoscopy. However, colonoscopies in persons with spinal cord injury are not without complications. We tested the hypothesis that, despite a high incidence of anemia, performing a colonoscopy in persons with spinal cord injury has a high complication rate and low rate of finding high risk pathology.

**Design:** Retrospective chart review

**Subjects:** 41 persons with chronic spinal cord injury admitted for colonoscopy from 2019-2024.

**Methods:** Percent of complications and abnormal findings were calculated. A logistic regression model determined predictors of complications and abnormal findings.

**Results:** Anemia prevalence was 59.1% with a complication rate of 38.6% and 59 abnormal findings in 95.2% of CSPs (n=4 (6.8%), high-risk pathology). Persons with anemia had a higher risk of complications and a decreased risk of hemorrhoids.

**Conclusion:** In persons with spinal cord injury, given a low-rate high risk pathology in the setting of a high complication rate, especially in persons with anemia, the risk of complications should be weighed more heavily in the decision to perform a colonoscopy.

## Introduction

As the second leading cause of cancer in the United States, the lifetime risk of colon cancer is about 1 in 23 for men and 1 in 25 for women, with an incidence rate of approximately 37.5 per 100,000 people. (1) For persons with spinal cord injury (SCI), the incidence of colon cancer appears comparable to persons without SCI, although the exact numbers can vary depending on factors like age, diet, and overall health.(2-4) Similarly, colorectal cancer mortality is also comparable to persons without SCI. (5)

Despite this comparable risk and incidence of colon cancer, performing colonoscopies (CSP) in individuals with SCI can present unique complications due to the underlying neurological and physiological changes caused by the injury. Autonomic dysfunction, impaired gastrointestinal motility, sensory deficits add a layer of medical complexity to this colon cancer screening procedure.(6) Often, a more prolonged bowel preparation is required in order to overcome the gastrointestinal dysmotility to adequately cleanse the colon prior to CSP. In addition to the added complexity of the preparation, peri procedural CSP complications in the SCI population are unique from the general population and at times requiring medical intervention, creating added procedural risk. Autonomic dysreflexia (AD), electrolyte imbalances, and pressure injuries have been cited as CSP related complications, however definitions of “complications” and methodology vary.(6-9) Further, data on the incidence of CSP procedure-related complications in relation to diagnosis of pathological findings in the SCI population remains limited. While the incidence of the most common CSP related complications is ≤1% (bleeding and perforation) in the general population, the incidence in persons with SCI is unknown (10)

This study was designed to measure 1) the incidence of peri-CSP complications in persons with SCI, 2) incidence of diagnosis of “high risk” pathology (e.g. gastrointestinal (GI) bleed, pre-cancerous polyp or cancerous lesion); 3) the association of a pre-procedure diagnosis of anemia with a CSP diagnosis of “high risk” or “low risk” (hemorrhoids and non pre-cancerous polyps) GI pathology and 4) the association of age, level of injury (tetraplegia vs. paraplegia), completeness of injury, time since injury, presence of anemia, incomplete bowel preparation or indication for CSP (diagnostic vs screening vs. surveillance) with CSP-complications. We hypothesize that in persons with SCI, the incidence of a peri-procedural CSP complication is greater than the general population while the diagnosis of “high risk” pathology is low altering the risk benefit ratio of CSP in persons with SCI.

## Methods

Following receipt of approval from local University affiliated Institutional Review Board and the local VA Research and Development board, retrospective chart review was conducted using the VA medical record (Computerized Patient Record System) of persons with SCI admitted to a Veterans Affairs (VA) SCI unit over a five-year period (2019-2024) for a CSP procedure.

Patients were included if they were at least one-year post-SCI with documented level of injury (defined by the International Standards for Neurological Classification after SCI exam) between C2-L2, and between the ages of 18 and 90 years old. Patients were excluded if they had a known source of bleeding or anemia (such as B12 deficiency), or if their CSP was indicated for surgical planning.

At this VA medical center, persons with SCI undergoing CSP were admitted 3 days prior for a bowel preparation if they were unable to physically undergo the bowel preparation at home safely. The bowel prep consisted of one gallon of polyethylene glycol 3350 (an osmotic laxative) on day 1, a second gallon of polyethylene glycol 3350 on day 2 and an enema 1 hour prior to CSP procedure on day 3.

The following information was collected: patient demographic data (age, time since injury), hemoglobin and hematocrit levels averaged over year prior, documented International Standards to document Neurological Classification after Spinal cord Injury (ISNCSCI) level and completeness, indication for CSP (i.e.; diagnostic vs. screening vs. surveillance), gross and histopathological findings and peri- or post-CSP complications requiring medical intervention up to 7 days post procedure. (11) Anemia was defined as hematocrit < 41%.

High-risk GI lesions included any of the following: GI bleed, carcinoma, tubulovillous adenoma, presence of high-grade dysplasia, serrated polyps, adenoma/villous adenoma > 10 mm, or 3 or greater adenomas of any size. Low risk lesions included less than 3 tubular adenomas < 10 mm, inflammatory polyps or hyperplastic polyps.

### Statistical Analysis

Primary outcome measures included 1) incidence of CSP related complications (with breakdown of each complication) and 2) Incidence of abnormal pathologic findings including rate of high-risk pathologic findings. Secondary outcomes included: 1) association between anemia and CSP findings of high-risk pathology, hemorrhoids or polyps and 2) association between CSP complication and age, level of injury (tetraplegia vs. paraplegia), completeness of injury, time since injury, anemia, incomplete bowel preparation or indication for CSP. A logistic regression model was used to calculate odds ratios (ORs) assessing the associations between all secondary outcome measures, with associated 95% confidence intervals and p-values.

## Results

Of the 68 charts reviewed for enrollment, 41 persons with SCI met inclusion and exclusion criteria. Of these 41, 3 persons had 2 colonoscopies over the 5-year period, for a total of 44 colonoscopies. Of the colonoscopies performed, the indications were as follows: 7 diagnostic, 21 screening, and 16 surveillance. The demographics of the study population are shown in Table I. Anemia was present in 24/41 patients (59.1%).

**TABLE 1.** Population demographics.

|  | n=41/CSP=44 |
| --- | --- |
| **Sex** |  |
| **Male** | 37 (90%) |
| **Female** | 4 (10%) |
| **SCI classification** |  |
| **Tetraplegia** | 15 (37%) |
| **Paraplegia** | 26 (63%) |
| **SCI level** |  |
| **cervical** | 15(37%) |
| **T1- T6** | 10 (24%) |
| **T7 and below** | 16 (39%) |
| **SCI AIS** |  |
| **complete (A/B)** | 24 (59%) |
| **incomplete (C/D)** | 17 (41%) |
| **Age at CSP** | 64.8 (9.5) |
| **Years since SCI** | 22.8 (14.9) |
| **Anemia (Hct <41%)** |  |
| **yes** | 24 (59%) |
| **no** | 17 (41%) |
| **CSP Indication** |  |
| **Diagnostic** | 7 (16%) |
| **Screening** | 21 (48%) |
| **Surveillance** | 16 (36%) |
| **Diagnostic indication** |  |
| **iron def anemia** | 4 |
| **anal bleeding** | 1 |
| **autonomic dysreflexia** | 1 |
| **bloating/constipation** | 1 |
| **incomplete prep** | 13 (30%) |

### Primary outcomes

Complications occurred in 17/44 colonoscopies, a rate of 38.6%. Of these, electrolyte abnormalities were the most common (n=5), followed by autonomic dysreflexia (AD) (n=3), ileus (n=2), elevated blood pressure not related to AD (n=2), hypotension (n=2), and other (n=3; urinary retention, new rectal bleeding secondary to CSP preparation requiring transfusion, and thrombocytopenia requiring transfusion). (Figure 1)

**FIGURE 1.**
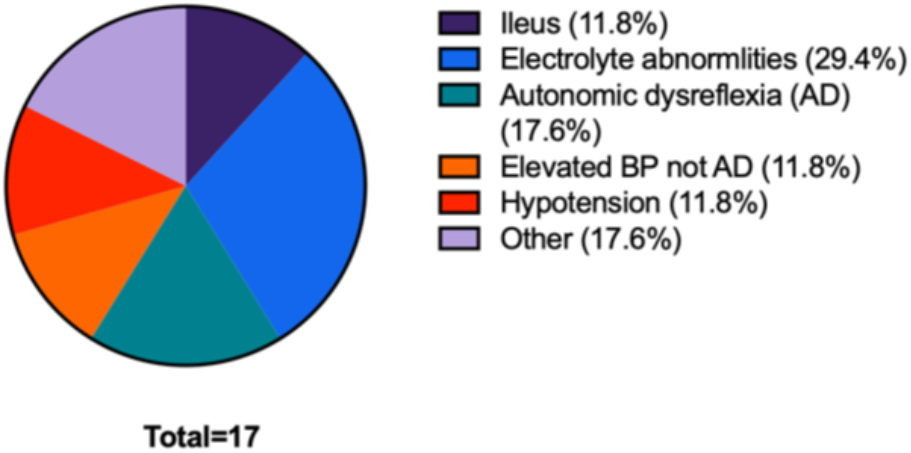
Breakdown of types of colonoscopy complications.

Abnormalities on CSP were found in 96% of (42/44) colonoscopies performed, totaling 59 lesions. Of these 59, *only 4 (6*.*8% of all lesions and 9*.*1% of all colonoscopies) “high-risk”* colonic lesions were found that pathology diagnosed as polyps ≥10mm in size. Of the remaining fifty-five “low risk” abnormalities found were: 30 polyps, 13 hemorrhoids, and 12 diverticuli. (Figure 2). There were no GI bleeds, carcinomas, tubulovillous adenomas, high grade dysplasia, serrated polyps or >3 lesions of any size found in any of the colonoscopies.

**FIGURE 2:**
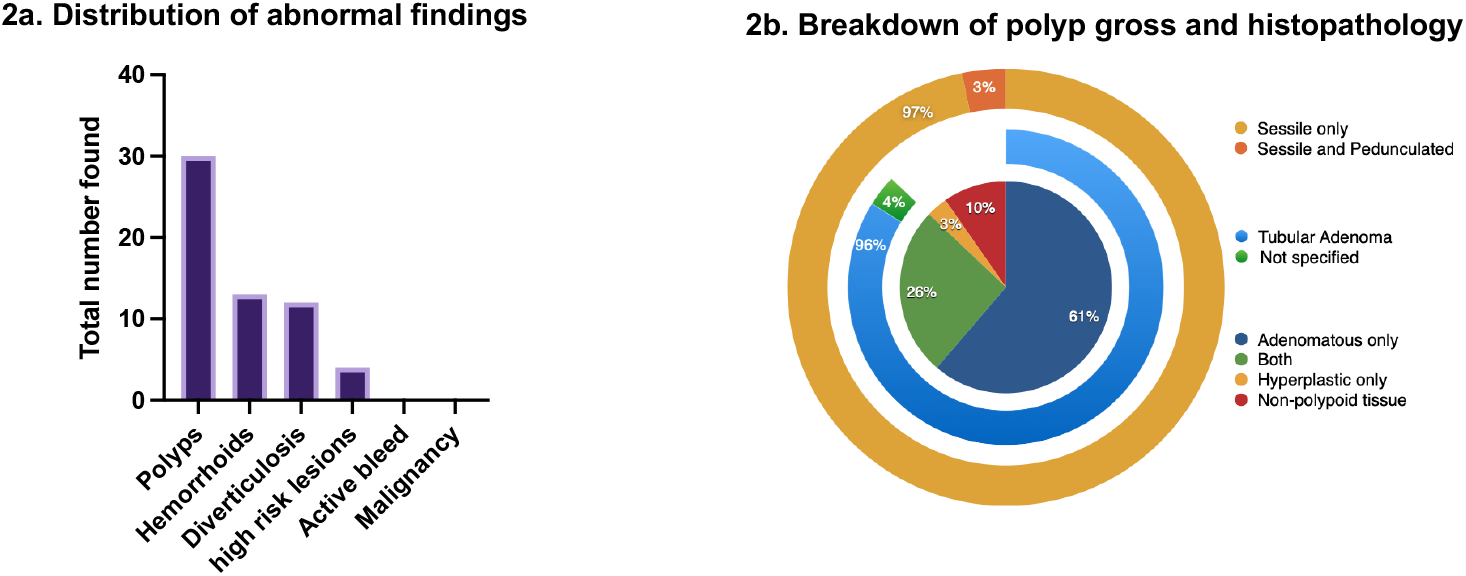
Distribution of a)abnormal findings and b) polyp gross and histopathology

### Secondary outcomes

Anemia was not a predictor of high-risk pathology of polyps >10mm (with OR 2.6 95% CI: [ 0.285 to 55.84], p=0.50). However, a statistically significant association was found between anemia and CSP-related complication, with an OR of 18 (95% CI: [2.8 to 234], p=0.008). (Table II) There was no statistically significant association between colonoscopy complications and age, level of injury, completeness of injury, time since injury, incomplete preparation, or indication for CSP. (Table II) Patients with anemia had statistically significant decreased odds of hemorrhoids, evidenced by OR 0.15 (95% CI: [0.032 to 0.55], p=0.007); there was no statistically significant relationship between anemia and polyps, OR 0.68 (95% CI: [0.13 to 3.1], p=0.624).

**TABLE 2:**
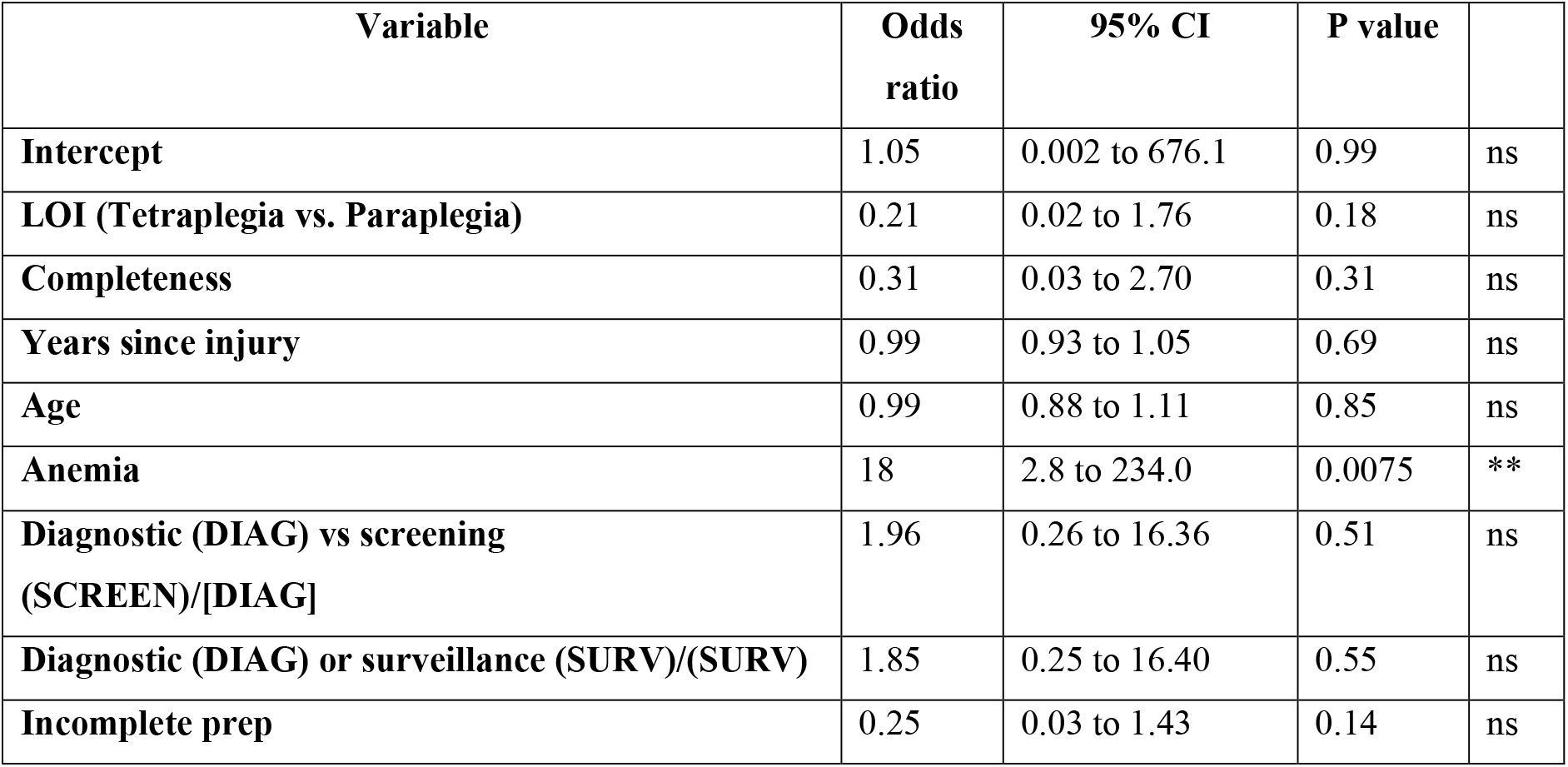
Multiple Logistical Regression Analysis of predictors of colonoscopy complications.

## Discussion

We found the incidence of CSP-related complications was 38.6% which is significantly higher than the incidence of CSP related complications in persons without SCI (≤1%). Further, this high complication rate was associated with a low incidence of “high-risk” colon pathology (9% of CSP/6.8% all pathology) consisting of polyps >10mm. There were no GI bleeds or cancerous lesions amongst the 44 CSP (59 abnormal lesions). Unexpectedly, persons with anemia had an 18-fold increased likelihood of experiencing a CSP-related complication. Interestingly we found that persons with anemia were: 1) not likely to have “high-risk” GI pathology; and 2) significantly less likely to have hemorrhoids. Age, level and completeness of injury, time since injury, incomplete preparation, or indication for CSP did not predict CSP complications.

### Incidence of complications post SCI

A large study found increased peri-colonoscopy complications in SCI patients compared to able-bodied, with rates of 4.7% vs 1.0%, respectively.(6) Another study followed 52 persons with SCI undergoing a non-traditional *extended* preparation (4 days with magnesium citrate and polyethylene glycol) and reported AD (n=5), electrolyte imbalances (n=18), and a pressure injury (n=1) for a 45.5% rate of complications.(7) We found a similar pattern of electrolyte imbalances as the most frequent complication, followed by AD with a total incidence of all complications at 39% in the setting of an 11% incomplete bowel cleansing, comparable to others.(7) AD during bowel preparation is reported in multiple case reports resulting from colorectal distension in persons prone to AD.(7-9)

Anemia was the only significant predictor of peri-CSP complication. While the explanation for this is unclear, we hypothesize that the increased likelihood of complications is due to cardiovascular compensation for chronic anemia (tachycardia and increased cardiac output) contributing to the blood pressure dysregulation complications (e.g.; AD, hypertension, hypotension) composing >50% (9/17) of the total complications post CSP. Further studies are needed to explore this unexpected result.

### Incidence of diagnosis of high vs. low-risk pathology

A 2012 study of 368 CSPs in 311 persons with SCI, 40% of CSPs resulted in polypectomies of 96 adenomas and 5 carcinomas for a rate of 27.4% of finding “high-risk” pathology.(12) This is higher than our finding of a 6.8% incidence of high-risk pathology in our smaller population (n=44, CSP). Meanwhile, Teng et al. found no difference in adenoma detection rate (55 vs. 51%, p = 0.59) or polyp histopathology (p = 0.748) between SCI and able-bodied populations.(4) Thus, it remains unclear if the risk of finding high-risk pathology in persons with SCI is comparable to able-bodied persons.

#### Association of anemia with pathology

Anemia prevalence in persons with chronic SCI is higher than their able-bodied counterparts, ranging from 52-88%.(13, 14) One review of 322 persons with chronic SCI, reported 72% were anemic (Hct≤40%) with only 7% being iron deficient.(15)

Similarly, our cohort fell within this range (∼60%). Notably, we found only 4/24 persons had iron deficiency anemia. The remaining 20/24 had normal MCVs consistent with anemia of chronic disease.

In our analysis, anemia was not a predictor of high-risk polyps >10mm. For low-risk pathology, the decreased odds of hemorrhoids and the non-significant association of polyps when anemia was present was unexpected and without obvious explanation. It does, however, suggest that the presence of hemorrhoids and polyps are *unlikely* to be the predominant explanation for anemia in chronic SCI persons.

In summary, our study indicates that physicians should consider and treat anemia from non-GI sources, prior to ordering a colonoscopy. Furthermore, considering the high complication rate, especially for the smaller cohort with anemia, it is worth considering noninvasive evaluation of GI sources, i.e., CT colonography, fecal immunochemical test or Cologuard as less risky alternatives.

#### Limitations

One limitation of this study is its design as a retrospective chart review. Furthermore, this study did not assess for increase in CSP surveillance frequency as an outcome of CSP findings, which could theoretically improve patient outcomes. While the presence of anemia was associated with increased odds of complications, we did not evaluate other risk factors (e.g.; medical comorbidities) that could more thoroughly elucidate CSP risks in SCI.

## Conclusion

In conclusion, given colon cancer is the 2^nd^ most common cancer, it is essential screening and diagnostic CSP are performed as recommend by Center for Disease Control guidelines.

However, persons with SCI have a higher incidence of complications compared to their able-bodied counterparts. SCI providers need to be aware of and inform their patients of the increased risk of complications including electrolyte abnormalities, autonomic dysreflexia, pressure sores, gastrointestinal complications, and blood pressure dysregulation. Further, subgroup of persons with SCI *and* anemia are more likely to have complications compared to persons without anemia so risks should be weighed more heavily during the consent process and non-invasive testing might be considered. Finally, pre-procedure assessment and preparation, along with clinical monitoring during and after the CSP procedure, are essential to minimize and mitigate these risks.

## Data Availability

All data produced in the present study are available upon reasonable request to the authors

## Ethics approvals

All human protocols were approved by the university affiliated Institutional Review Board and therefore were performed in accordance with the ethical standards laid down in the 1964 Declaration of Helsinki and its later amendments. Veterans Affairs local Research Development office approved all protocols in this study. All participants provided written informed consent to participate.

